# ReMiND: Recovery of Missing Neuroimaging using Diffusion Models with Application to Alzheimer’s Disease

**DOI:** 10.1101/2023.08.16.23294169

**Authors:** Chenxi Yuan, Jinhao Duan, Nicholas J. Tustison, Kaidi Xu, Rebecca A. Hubbard, Kristin A. Linn

## Abstract

**Objective:** Missing data is a significant challenge in medical research. In longitudinal studies of Alzheimer’s disease (AD) where structural magnetic resonance imaging (MRI) is collected from individuals at multiple time points, participants may miss a study visit or drop out. Additionally, technical issues such as participant motion in the scanner may result in unusable imaging data at designated visits. Such missing data may hinder the development of high-quality imaging-based biomarkers. Furthermore, when imaging data are unavailable in clinical practice, patients may not benefit from effective application of biomarkers for disease diagnosis and monitoring.

**Methods:** To address the problem of missing MRI data in studies of AD, we introduced a novel 3D diffusion model specifically designed for imputing missing structural MRI (<u>R</u>ecovery of <u>M</u>issing <u>N</u>euroimaging using <u>D</u>iffusion models (ReMiND)). The model generates a whole-brain image conditional on a single structural MRI observed at a past visit or conditional on one past and one future observed structural MRI relative to the missing observation.

**Results:** Experimental results show that our method can generate highquality individual 3D structural MRI with high similarity to ground truth, observed images. Additionally, images generated using ReMiND exhibit relatively lower error rates and more accurately estimated rates of atrophy over time in important anatomical brain regions compared with two alternative imputation approaches: forward filling and image generation using variational autoencoders.

**Conclusion:** Our 3D diffusion model can impute missing structural MRI data at a single designated visit and outperforms alternative methods for imputing whole-brain images that are missing from longitudinal trajectories.

## 1. INTRODUCTION

Alzheimer’s disease (AD) is a progressive neurodegenerative disorder characterized by a decline in cognitive abilities, including memory, language and problem-solving abilities [1]. The accurate prediction of progression from normal cognition to mild cognitive impairment (MCI) and subsequently to AD will become increasingly important for patient care and resource allocation as early interventions and treatments for the disease are developed [2]. The diagnosis of AD involves a variety of modalities, including clinical evaluations, neuropsychological testing, biomarker analysis, and brain imaging [2, 3, 4]. Brain imaging techniques, such as magnetic resonance imaging (MRI), can provide information about changes in brain structure or function that occur as AD progresses [5]. MRI feature-based classification and prediction algorithms have a high potential for early detection of characteristic AD patterns in brain structure and activity [6]. In research studies that use repeated longitudinal imaging to measure brain changes over time, planned imaging scans may be missing due to participant dropout, technical issues during image acquisition, or participant unwillingness to undergo imaging, resulting in the absence or incompleteness of imaging data trajectories for some study participants [7, 8]. The study’s validity and power can both be significantly impacted by the effects of missing imaging data.

Longitudinal studies frequently suffer from missing data due to the multiple rounds of data collection over time that increase the chance of nonresponse and participant attrition [9]. In studies of older adults, there is a high risk of missing data due to the susceptibility of this population to physical and cognitive decline, illness, and death [10], which may impact completion of assessments. The presence of missing data poses several challenges for longitudinal studies of AD, such as reducing the sample size overall or disproportionately in the AD-affected group, introducing selection bias, and reducing statistical power for estimating and evaluating the effect of imaging biomarkers [11]. Researchers in AD have made efforts to impute longitudinal missing data by applying various techniques, such as forward filling, linear filling, K-Nearest Neighbor, multiple kernel learning, and recurrent neural networks [7, 12, 13, 14, 15]. However, the majority of existing methods generated image-derived phenotypes (IDPs) rather than imputing the entire missing image. In this work, we employ a diffusion model to generate an entire 3D image conditional on one or more observed images from an individual’s imaging trajectory.

The denoising diffusion probabilistic model (DDPM or diffusion model for short) [16], is a new class of generative models that utilizes a latent variable framework to reverse a diffusion process, wherein Gaussian noise is gradually added to alter the data distribution to the noise distribution. Diffusion models are applied to tasks such as image, audio, and graph production, as well as conditional generation tasks such as in-painting, super-resolution, and picture editing [17]. Diffusion models have demonstrated exceptional performance in various tasks [18, 19, 20, 17] and are well suited to our longitudinal imputation problem in three respects. First, the diffusion model shows promising results when synthesizing natural images and has rivaled state-of-the-art models such as generative adversarial nets and variational autoencoders [21, 22]. Second, diffusion models have more flexible condition configurations to create images conditional on other features [23, 24], while conventional generative models may require additional annotations. Third, the diffusion model has the ability to generate images in the temporal dimension, such as video-related tasks that predict the future frame conditioned on the past frame [25]. Longitudinal MRI image imputation is analogous to image generation in the temporal dimension. To address this, we have developed a novel approach called ReMiND (<u>Re</u>covery of <u>Mi</u>ssing <u>N</u>euroimaging with <u>D</u>iffusion models) for 3D MRI imputation in longitudinal studies of AD. ReMiND focuses on generating missing images at a designated single visit by conditioning on one or more observed images from other time points. The overall pipeline of the proposed ReMiND approach is illustrated in Fig. 1. The main contributions of this research are as follows:

**Figure 1.**
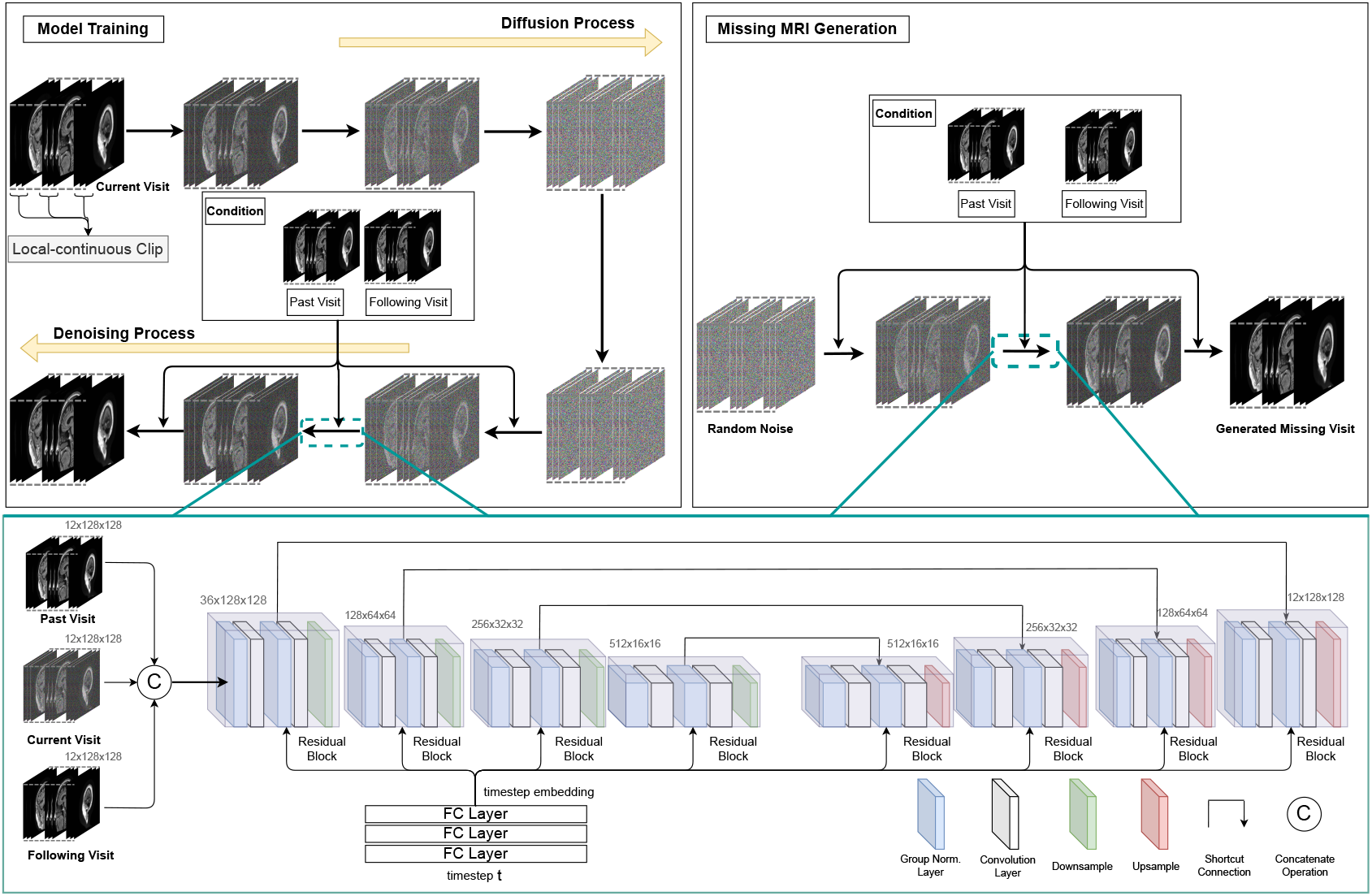
Pipeline of the proposed ReMiND model. For model training, ReMiND follows the diffusion process of a DDPM by adding random noise on a designated MRI. Then, ReMiND leverages parameterized neural networks during the denoising process to recover the noise applied, with conditions over past visits or past and following visits. To generate a missing MRI after the model is trained, ReMiND passes random noise through the learned denoising process along with one or more observed images from other time points in a subject’s longitudinal image trajectory. The denoising process is parameterized by UNet-like neural networks, taking the concatenation of conditions and intermediate results as input and predicting the added noise. FC Layer refers to Fully-Connected Layer. Group Norm. refers to Group Normalization, which first aggregates activations into groups by channels and then calculates group-wise normalization. Shortcut connection refers to a branch where the source of the connection will be directly added to the end, which benefits multi-scale modeling and gradient propagation. Residual block means there will be one shortcut connection within this block.

1. Unlike previous studies that utilized multiple imaging modalities to impute missing imaging data, our work focuses on imputing missing MRI images in the temporal dimension using images from the same modality at other time points, specifically in the context of AD.

2. We have developed a novel 3D diffusion model specifically designed for MRI image generation. The model effectively preserves global information of the whole MRI image through the incorporation of local-continuous slices. As a result, the model produces high-quality and plausible 3D structural MRIs.

3. The proposed model imputes the missing 3D MRI images directly rather than imputing 2D slices or image derived phenotypes (IDPs). The availability of imputed whole-brain MRI will allow researchers to derive any summary measures of choice using any software of choice without having to adapt or re-run an imputation procedure specific to a certain IDP or software pipeline.

4. The proposed model conditions on a limited set of images (either past or both past and following visits) to generate the missing image for each subject, which is specifically tailored for the analysis of longitudinal data.

These developments collectively contribute to the advancement of longitudinal MRI image imputation and analysis techniques for AD research.

## 2. METHODS

### 2.1 Denoising Diffusion Probabilistic Models (DDPM)

DDPM [16] is a form of latent variable model that approximates the real data distribution **x**_0_ *∼ p*_*data*_ with a diffusion process *q*(**x**_*t*_|**x**_*t−*1_), *t ∈{*1, …, *T }*, and a denoising process *p*_*θ*_(**x**_*t−*1_|**x**_*t*_) parameterized by weights *θ*. The diffusion process transmits *p*_*data*_ to a standard normal distribution with a *T* -step Markov chain:

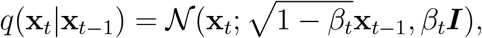

where the observed image **x**_0_ *∈* ℝ*d* is assumed to be a draw from *p*_*data*_ and *β*_1_, *β*_2_,, *β*_*T*_ is a fixed variance schedule. The forward sampling at arbitrary time step *t* is defined as

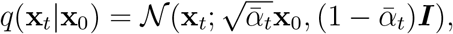

where *α*_*t*_ = 1 *−β*_*t*_ and 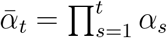 Then, a denoising process parameterized by weights *θ* is leveraged to match the diffusion process at each timestep *t* with the following transition kernel:

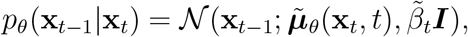

where

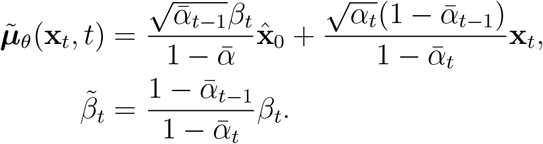

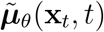 and 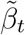 are the mean and the variance of the posterior distribution 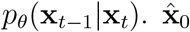 refers to the estimated **x**_0_ at timestep *t*

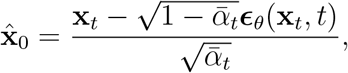

and ***ϵ***_*θ*_ is a neural network trained to predict noise ***ϵ***, e.g., the UNet [26]. The learning objective of DDPM is to optimize the variance bound of *p*_*θ*_(**x**_0_) which can be simplified as the “noise-prediction” loss as in [16]:

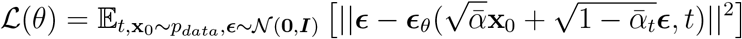

### 2.2 Recovery of Missing Neuroimaging with Diffusion models (ReMiND)

In this section, we formulate the longitudinal MRI imputation problem as a generation task conditioned on one or more adjacent MRI images of the designated missing visit. For a given subject ***S****∈ {*1, …, *N }*, we define the longitudinal record of ***S*** as a set of (Image, Existence) pairs arranged in order of visiting timepoint:

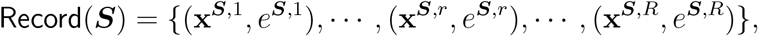

where *R* refers to the number of records contained in Record(***S***) if the data were fully observed, i.e., *R* = |Record(***S***)|, and **x**^***S***,*r*^ *∈* ℝ*L×H×W* refers to the 3D structural MRI image at the *r*-th visit with resolution *L× H× W*. *e*^***S***,*r*^ is the indicator of existence, i.e., *e*^***S***,*r*^ = 0 means **x**^***S***,*r*^ is missing; *e*^***S***,*r*^ = 1 means **x**^***S***,*r*^ exists in the data. Without loss of generality, we assume the *r*-th image is missing, i.e., *e*^***S***,*r*^ = 0, and its adjacent visits exist, i.e., *e*^***S***,*r−*1^ = *e*^***S***,*r*+1^ = 1. We impute **x**^***S***,*r*^ by taking its adjacent neighbors as conditions:

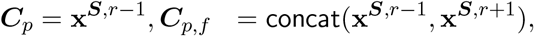

where ***C***_*p*_ refers to the condition over the past visit and ***C***_*p,f*_ refers to the condition over both the past visit and the following visit. concat(*·,·*) is the concatenate operation.

Since we aim to recover missing images from neighboring visits where imaging is available, we formulate the longitudinal imputation method as a conditional image generation task. Specifically, ReMiND aims to approximate the distribution of a missing image, 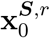 with a parameterized conditional distribution, 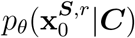 In the form of diffusion and denoising transitions, ReMiND matches 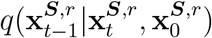 with 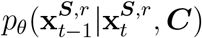 at each timestep *t*, where 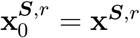 and ***C*** = ***C***_*p*_ or ***C***_*p,f*_.

To achieve this, the denoising process is re-written as:

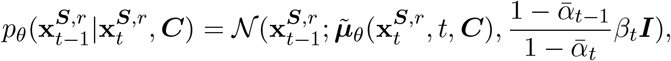

where

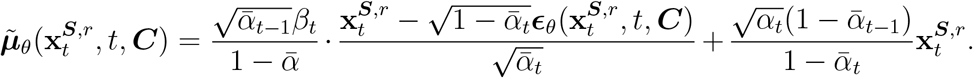

The diffusion process of ReMiND is the same as a DDPM except we replace the desired data distribution with 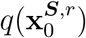 In this way, the learning objective of ReMiND is:

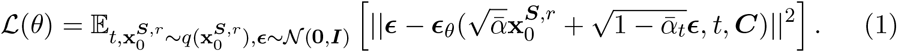

Henceforth, we denote the model that only relies on the *past* visit as ReMiNDP, i.e., ***C*** = ***C***_*p*_, and the model that relies on both the *past* visit and the *following* visit as ReMiND-PF, i.e., ***C*** = ***C***_*p,f*_.

During training, Eq. (1) requires that **x**^***S***,*r*^ be recovered from random noise if the condition ***C*** is given during the denoising process at each timestep. Once the ReMiND model has converged over Eq. (1), ***ϵ***_*θ*_ will capture the spatial-temporal dynamics across the missing image and its longitudinal neighboring images. Therefore, for imputation, as long as the condition is provided as prior information, the denoising process will gradually generate the desired missing image. In this work, we only consider models that condition on the immediate adjacent visits, i.e., ***C***_*p*_ and ***C***_*p,f*_, since these two conditions represent two commonly encountered missingness patterns in real longitudinal data. Our method can also easily be generalized to impute missing visits that occur within longer visit trajectories, i.e., multiple past/following visits as conditions.

Normally, modeling high-resolution 3D structural MRIs requires highcapacity 3D convolutional neural networks [27, 28]. However, these models are computationally intensive. For instance, the resolution of MRI used in our application is 256*×* 256*×* 172 voxels, which requires enormous GPU memory for training and is generally computationally infeasible for such models. Furthermore, high-capacity models are known to generalize poorly on smallscale datasets such as those available from AD research studies where the number of study participants and images per participant are limited [29]. Although 2D convolutional networks are likely computationally feasible for AD applications, they can only involve at most two dimensions during computing, which leads to locally non-continuous 3D MRI generations.

In this paper, we mitigate these issues via a parameter-efficient training paradigm by splitting 3D MRIs into uniform local-continuous clips and training 2D convolutional neural networks over these clips. Concretely, for a given 3D structural MRI with the shape of **x***∈* ℝ*L×H×W*, it has length *L*, width *W*, and height *H*. We split **x** into *K* segments uniformly along the *L*-axis. Each segment has a resolution of ^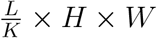^ During model training, we first randomly select one image slice with shape *H× W*, from each segment. Then, we concatenate each selected slice with *J* slices immediately before it and *J* slices immediately after it as the local-continuous clip, which achieves the shape of *K*(2*J* + 1) × *H ×W* for each clip. These clips are the basic units for model training and only one clip will be fed into the model at each optimization step. In other words, instead of randomly selecting slices from segments, we construct local-continuous clips by selecting three consecutive slices (i.e., we use J=1) from each segment. Theoretically, there will be *K*(2*J* + 1) slices for each clip and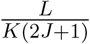clips for each 3D MRI. We feed these 3D clips into the model and finally reassemble the outputs into the 3D MRI with the original shape, i.e., *L× H ×W*.

The reason we construct local-continuous clips in this way is two-fold: 1) slices within each clip are uniformly drawn from all segments across the entire *L*-axis. It indicates that all clips encompass the global information of the whole 3D MRI, which aids the completeness of generations; 2) combining slices with their immediate neighbors preserves local information and overcomes the insufficient modeling of 2D convolutional networks. In this way, our method yields smooth and continuous 3D images.

## 3. EXPERIMENTS

### 3.1 Dataset and Preprocessing

To illustrate the utility of ReMiND, we leverage longitudinal MRI data that are publicly available from the Alzheimer’s Disease Neuroimaging Initiative (ADNI) [30], which was initiated in 2003 with the goal of facilitating the study of AD. In brief, ADNI enrolled participants between the ages of 55 and 90 who were recruited at 57 sites in the United States and Canada. The dataset we use comprises T1-weighted MRI from participants who provided data to ADNI on at least two separate visits between September 2005 and May 2017, with a fixed interval of 6 months between each visit. The sample sizes of each clinical diagnosis that we used for training, validation, and testing are presented in Table 1.

**Table 1.**
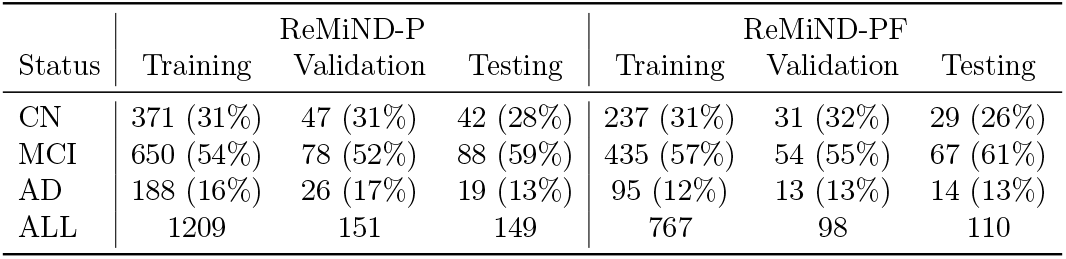
Distribution of observations used for ReMiND-P and ReMiND-PF models overall and stratified by clinical status including cognitively normal (CN), mild cognitive impairment (MCI), and Alzheimer’s disease (AD). Each value is the sample size followed by the percentage of total observations in the corresponding column.

| Status | ReMiND-P |  |  | ReMiND-PF |  |  |
| --- | --- | --- | --- | --- | --- | --- |
|  | Training | Validation | Testing | Training | Validation | Testing |
| CN | 371 (31%) | 47 (31%) | 42 (28%) | 237 (31%) | 31 (32%) | 29 (26%) |
| MCI | 650 (54%) | 78 (52%) | 88 (59%) | 435 (57%) | 54 (55%) | 67 (61%) |
| AD | 188 (16%) | 26 (17%) | 19 (13%) | 95 (12%) | 13 (13%) | 14 (13%) |
| ALL | 1209 | 151 | 149 | 767 | 98 | 110 |

We pre-processed the T1-weighted images by following the ANTs longitudinal cortical thickness pipeline [31]. For each subject, we first built a single-subject template (SST) using all longitudinal images belonging to that subject followed by rigid registration of each image into the SST space. Next, we rigidly registered each SST to a global template and aligned all within-subject images to the global template by applying the warps from the corresponding SST registration. To reduce computational costs, we rescaled each axial slice from 256×256 to 128×128 voxels, resulting in 170 *×*128*×* 128 resolution for each image. Finally, we applied min-max normalization to each image. Since the background voxels in each image have a value of 0, the normalization procedure was considered to be applied exclusively to the voxels within the skull/brain region.

### 3.2 Experiment Setting

We used T1-weighted MRI from 632 ADNI participants with clinical status classified as: cognitively normal (CN), mild cognitive impairment (MCI), or AD. We performed 10-fold cross-validation for model selection and evaluation. After randomly partitioning the data into 10 equal subsets, each iteration of the 10-fold cross-validation utilized 80% for model training, 10% for model validation, and the remaining 10% for testing. In each iteration, the training set was used for model fitting, the validation set was used to select values for hyperparameters, and the test set was used to evaluate the model’s performance under the optimal set of hyperparameters identified by the validation set. We trained separate imputation models for two settings: (1) the ReMiND-P model imputes a missing image given the most recent past visit, and (2) the ReMiND-PF model imputes a missing image given the most recent past visit and the future visit that follows the missing time point. We simulated missingness in the dataset for this study by manually selecting some visits from the complete data. Specifically, for the ReMiND-P model, every second visit was considered as a missing data point. On the other hand, for the ReMiND-PF model, the middle visit was regarded as missing data among every three observed timepoints. We refer to the selected missing image, which actually exists in the dataset, as the “observed image” in this study.

To assess the performance of the ReMiND models, we first calculated the structural similarity index (SSIM) and the peak signal-to-noise ratio (PSNR) to quantify the proximity of the imputed images to the observed images. SSIM is an algorithm that checks the similarity between two images based on three factors: luminance, contrast, and structure. It is designed to better suit the human visual system and capture perceptual changes in the image. SSIM ranges from -1 to 1, where 1 means perfect similarity [32]. PSNR is a ratio that measures the amount of noise or distortion introduced by compression or reconstruction. It is based on the mean squared error between the two images. The higher the PSNR, the better the quality of the image [33]. We further compared regional brain volumes estimated using two common pipelines: 1) the ANTs longitudinal cortical thickness pipeline [31] and 2) FreeSurfer [34]. We employed the error rate and progression rate as metrics to facilitate the comparison. The error rate, calculated as |ŷ_*i*_ *− y*_*i*_|*/y*_*i*_, compares the volume estimation of a specific region from the imputed imageŷ_*i*_ to that from the observed image *y*_*i*_. Lower error rate indicates better performance. The progression rate, on the other hand, measures the rate of volume decline, reflecting brain atrophy between adjacent visits. For imputed images, the progression rate is computed as |ŷ_*i*_ *− y*_*i−*1_|*/y*_*i−*1_. Here, *y*_*i−*1_ represents the volume of the previous visit, and |ŷ_*i*_*− y*_*i−*1_ |represents the change in brain volume in imputed images at a specific visit compared to the previous one.

Similarly, the progression rate for observed images is |*y*_*i*_*− y*_*i−*1_ |*/y*_*i−*1_. The smaller difference between the progression rate using an observed image and the progression rate using an imputed image signifies better performance.

Furthermore, we studied the relative performance of ReMiND versus two comparator models: naive imputation by forward filling (Naive) and imputation using an autoencoder (AE). The Naive-P model simply predicted all missing images to be the same as the past observed images. The Naive-PF model predicted the missing images by averaging the adjacent past and future images. AE models are widely used for image processing and generation [35, 36, 37, 38]. We trained AE models to take as input the past or past and following visits and then minimize the *ℓ*_2_ loss between the output (i.e., imputed image) and the target “missing” MRI. Thus, AE-P refers to taking the past visit as input and AE-PF refers to taking both the past visit and the following visit as input to impute the missing MRI. We additionally compared performance of each imputation method and processing pipeline separately by clinical diagnosis group. However, training utilized data pooled from all groups.

### 3.3 Implementation Details

We adopted a modified UNet [39] with larger capacity and additional attention blocks. Models were trained in 200,000 steps with Adam [40] as the optimizer. We followed the UNet architecture described in [16] except for the model size, where we adopted the channel multiplier as 64 for ReMiND-P and 128 for ReMiND-PF since ReMiND-PF consists of larger conditions. We trained both ReMiND-P and ReMiND-PF in 200,000 steps with the Adam optimizer [40]. The learning rate was set to 0.0001 and the batch size was set to 16 during the training. We adopted the same U-Net architecture as ReMiND for the AE models. Since the AE models tend to converge easily, we trained the AEs until the loss stopped decreasing (around 20,000 steps) with the learning rate set as 1e-4. All experiments were conducted on servers consisting of one Nvidia RTX 3090 GPU, one Intel i9-12900F CPU, and 32G RAM. PyTorch [41] was used as the deep learning framework in our implementation.

## 4. RESULTS

### 4.1 Whole-Brain Imputed Images

To qualitatively evaluate the ReMiND-generated images, we visually compared the imputed images with their corresponding observed images for one subject in Fig. 2. The presented images were generated with the models conditioned on both past and following images, as ReMiND-PF outperformed the ReMiND-P model on the quantitative measures which we will report next. To visually demonstrate the superior performance of the ReMiND-PF method, we display corresponding slices from the images generated using the Naive-PF and AE-PF models. For all methods, images were intentionally generated with the skull on to prioritize flexibility in downstream analyses. That is, researchers working with the imputed image could apply their brain extraction method of choice.

**Figure 2.**
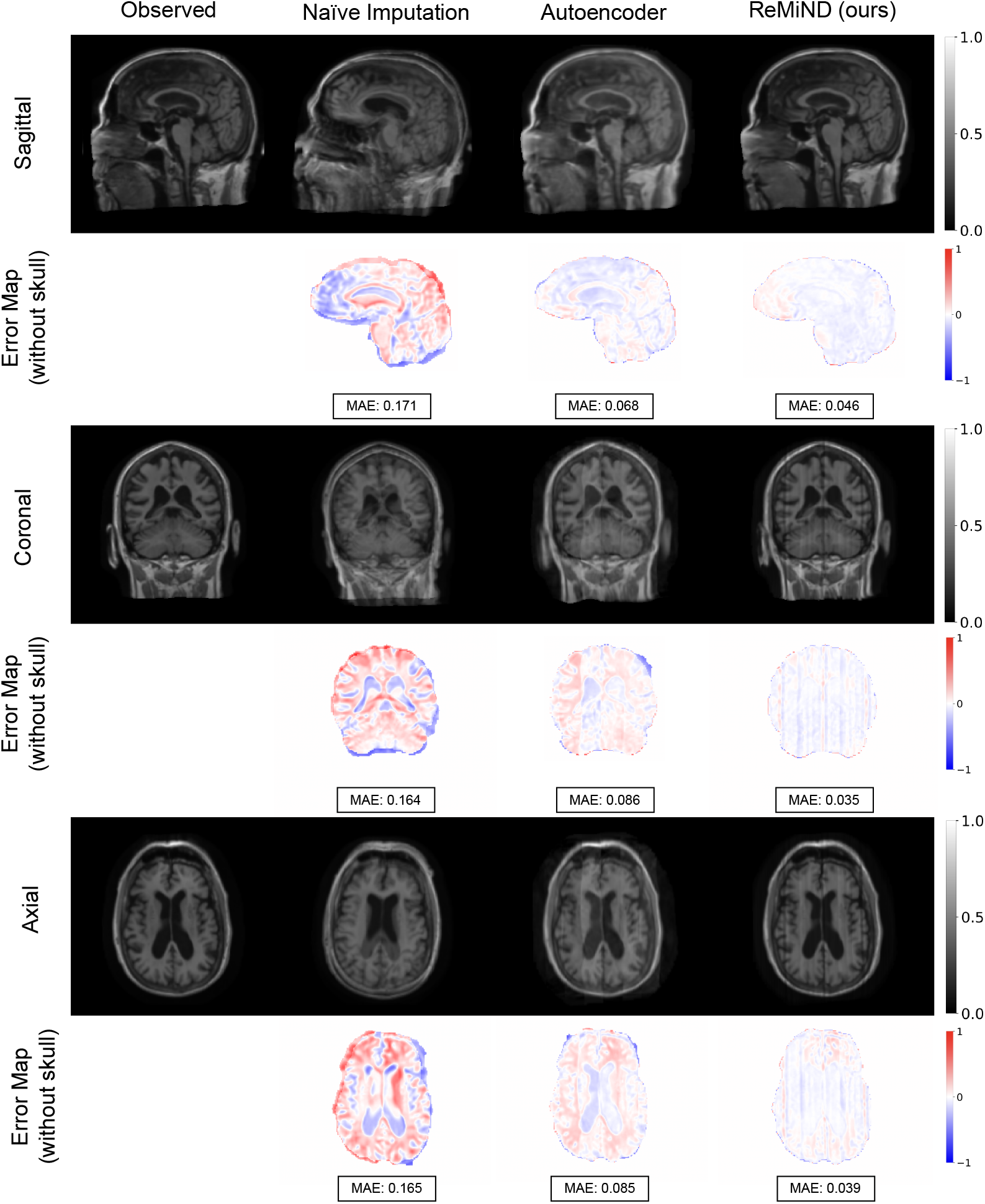
Qualitative comparison of observed images and imputed T1-weighted MRI which were generated by Naive-PF, AE-PF, and the proposed ReMiND-PF models. The first, third, and fifth rows show the generated images from sagittal, coronal, and axial views, respectively. The second, fourth, and last row shows the error maps of brain voxelwise differences between the observed and generated images. MAE indicates the mean absolute error.

As shown in Fig. 2, images generated with the ReMiND model are visually more similar to the observed images compared to the other two imputation methods. The Naive-PF imputed images have several blurry areas and imprecise skulls due to averaging across rigidly-registered images from different visits. Images imputed using the autoencoder model are marginally better than those generated using the na ï ve approach but still exhibit undesirable artifacts and fuzzy edges. Compared to the other methods, the ReMiND model generated images with sharper edges and finer anatomical details in critical gray matter regions for this individual. Based on the qualitative comparison in Fig. 2, our method appears to capture important anatomical structures such as the cortical gray matter with high integrity. The differing performance of the three imputation methods is further highlighted by the error images of brain voxel-wise differences between the generated and observed images. The Naive imputation exhibited the largest dissimilarity between observed and imputed images, while ReMiND preserved fine structural details resulting in small voxelwise differences across the brain.

Table 2 quantifies the proximity of the imputed images to the observed images at the target visit, considering images with skull voxels included. Across all clinical statuses, ReMiND models outperformed the Naive and AE models with respect to SSIM and PSNR values. This finding held both when imputing conditional on the past image and conditional on the past and following images. The ReMiND-PF model had both larger SSIM and PSNR than the ReMiND-P model, suggesting the model performs better when more than one time point is available to condition on for imputation. In addition to testing the models on the whole testing dataset, we evaluated the performance of all methods separately amongst groups of subjects categorized as CN, MCI, and AD. We found only minor differences between the all-status group and each clinical status group for both SSIM and PSNR. Our findings suggest that the methods considered can effectively impute missing structural MRI data for patients across the spectrum of clinical severity.

**Table 2.** Comparison of model performance averaged across 10 test sets. Performance was measured with structural similarity index (SSIM) and peak signal-to-noise ratio (PSNR) in decibels (dB) on the generated MRI images with skull voxels included in the calculation. Performance was evaluated overall and separately by clinical group (CN, MCI, and AD). P indicates the imputation method conditioned on the most recent past image. PF indicates the imputation method conditioned on both the most recent past image and the closest following image. AE indicates imputation using an autoencoder. Bold values indicate the best performing imputation method for a given clinical diagnosis group within the P or PF condition.

| Model | Images | CN | MCI | AD | ALL |
| --- | --- | --- | --- | --- | --- |
| SSIM (higher=better) |  |  |  |  |  |
| Naive-P | w/ skull | 0.705 ± 0.032 | 0.710 ± 0.013 | 0.747 ± 0.026 | 0.714 ± 0.015 |
| AE-P | w/ skull | 0.730 ± 0.010 | 0.725 ± 0.015 | 0.735 ± 0.017 | 0.728 ± 0.011 |
| ReMiND-P | w/ skull | **0.850 ± 0.010** | **0.848 ± 0.006** | **0.850 ± 0.011** | **0.850 ± 0.007** |
| Naive-PF | w/ skull | 0.702 ± 0.061 | 0.695 ± 0.026 | 0.707 ± 0.016 | 0.701 ± 0.014 |
| AE-PF | w/ skull | 0.728 ± 0.006 | 0.739 ± 0.018 | 0.746 ± 0.016 | 0.737 ± 0.013 |
| ReMiND-PF | w/ skull | **0.886 ± 0.010** | **0.899 ± 0.010** | **0.900 ± 0.004** | **0.895 ± 0.002** |
| PSNR (dB) (higher=better) |  |  |  |  |  |
| Naive-P | w/ skull | 22.169 ± 1.253 | 22.364 ± 0.551 | 23.704 ± 1.413 | 22.503 ± 0.692 |
| AE-P | w/ skull | 25.013 ± 0.274 | 24.630 ± 0.442 | 24.837 ± 0.617 | 24.780 ± 0.319 |
| ReMiND-P | w/ skull | **27.182 ± 0.549** | **26.939 ± 0.171** | **27.455 ± 0.966** | **27.101 ± 0.285** |
| Naive-PF | w/ skull | 22.241 ± 2.691 | 22.112 ± 0.851 | 22.267 ± 0.280 | 22.255 ± 0.682 |
| AE-PF | w/ skull | 24.805 ± 0.227 | 24.907 ± 0.343 | 25.002 ± 0.139 | 24.890 ± 0.273 |
| ReMiND-PF | w/ skull | **29.192 ± 0.608** | **28.870 ± 0.455** | **28.819 ± 0.833** | **28.956 ± 0.406** |

The results for images without the skull are provided in supplementary material Table A.5. SSIM and PSNR values were higher when computed on brain images without skull voxels compared to images with the skull included. This is likely due to across-subject heterogeneity in extra-cerebral voxels that display the neck and facial features. Since extra-cerebral regions are not important for studying the effects of AD in the brain over time, evaluation of ReMiND and other methods should focus on metrics with the skull removed.

### 4.2 Evaluation of Volumetric Features Extracted from Generated Images

We evaluated the performance of the proposed ReMiND models with respect to volumetric features extracted using the longitudinal ANTs volume-based cortical thickness estimation pipeline[42] and FreeSurfer’s pipeline [34]. Results are presented in Table 3. We primarily focused on 28 cortical regions that have been shown to be associated with AD pathology in the brain. Detailed region names are provided in the supplementary material Table A.4. Results in Table 3 are based on the average across these 28 regions and are also averaged across the 10 testing sets.

**Table 3.**
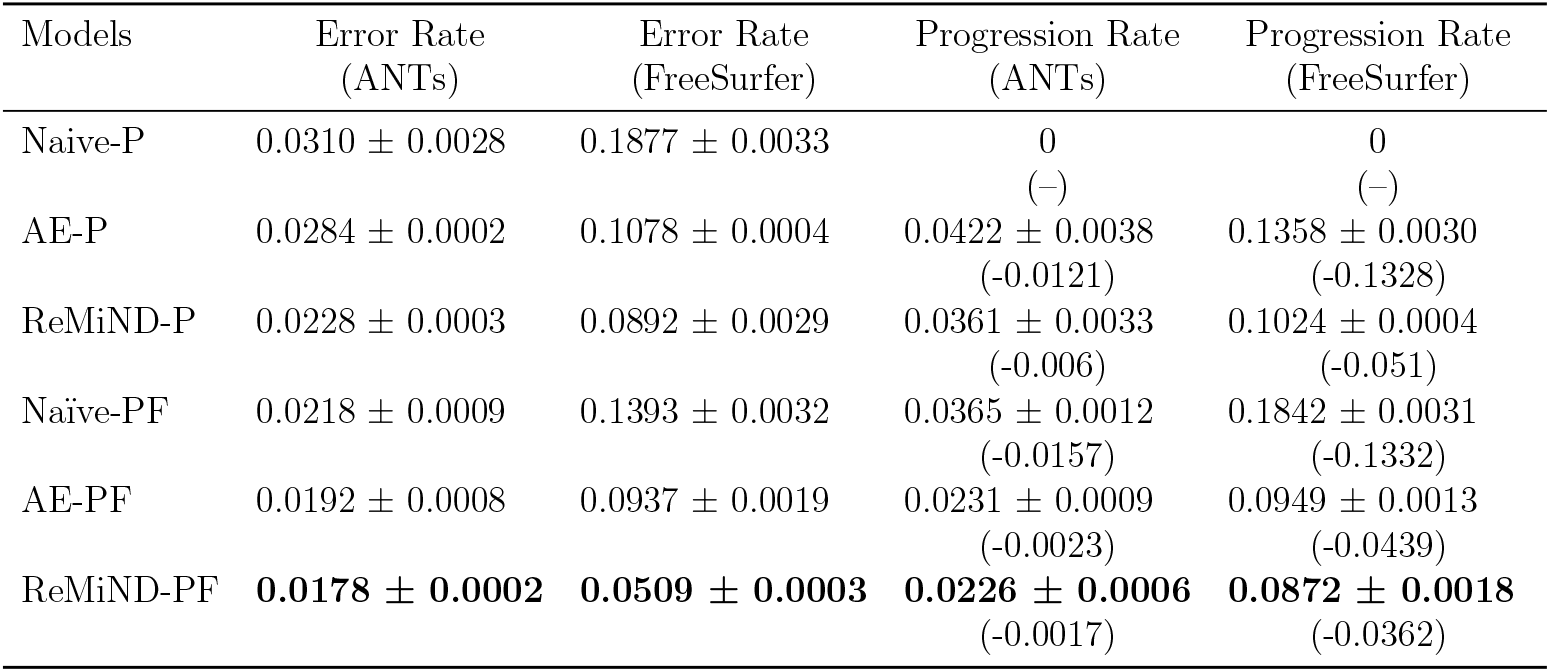
Comparison of the test performance of imputation methods with respect to volumetric features extracted from whole-brain imputed images. Results were averaged across the 10 test sets and averaged across all cortical regions defined by the DesikanKilliany-Tourville atla. Lower error rate indicates better performance. The values in () are differences between the observed progression rate (using observed images at both time points) and the estimated progression rate using the imputed image at the latter time point. Lower difference means better performance. P indicates past image. PF indicates past and following images. The best test results across all methods and models are bolded.

We found the error rates were lower for PF models compared to P models across all three methods (Naive, AE, and ReMiND). These results suggest models that condition on past and following images perform better at the task of generating accurate missing MRI images at the designated visits, which is likely explained by the additional information of the subsequent observed image. Under each experiment setting (P and PF), ReMiND models have the lowest error rates compared with two comparator methods, and the Naive method had the highest error rates. Furthermore, ANTs-based error rates were lower across all methods and experimental settings compared to FreeSurfer which is an expected finding based on previous work [42].

The progression rates for Naive-P model are all zero in Table 3 because the Naive-P model generates the missing image simply by carrying forward the past image. The ReMiND-P model had lower differences in observed and imputed progression than AE-P using both the ANTs and FreeSurfer pipelines. Furthermore, ReMiND-PF models exhibited the lowest differences between the observed and imputed progression rates for both volume estimation pipelines. FreeSurfer-based differences were larger than ANTs-based differences in progression rates.

In addition to results averaged across all 28 prioritized brain regions, we report results from the hippocampus, parahippocampal region, and the third ventricle individually in Fig. 3. The figure displays total estimated volume (in mm^3^), error rate, and progression rate for all imputation methods and both P and PF models. All results were averaged across 10 test sets. Not surprisingly, the performance reflects the results in Table 3. ReMiND models outperform both comparator imputation approaches. Specifically, Fig. 3 (panels a-c) shows that ReMiND models exhibit smaller discrepancies between the estimated and observed values compared to Naive and AE models. Fig. 3 (panels d-f) demonstrates that ReMiND-generated images have the lowest error rates across methods, particularly when the FreeSurfer pipeline is used to extract the volumes. The third row of Fig. 3 (panels g-i) demonstrates that the ReMiND-P and ReMiND-PF models produce the smallest differences between imputed and observed progression rates across imputation methods. Although the estimated volumes may not exhibit significant visual distinctions, the observed differences in error rate and progression rates in the plot are primarily influenced by the difference between the estimated and observed values for each method.

**Figure 3.**
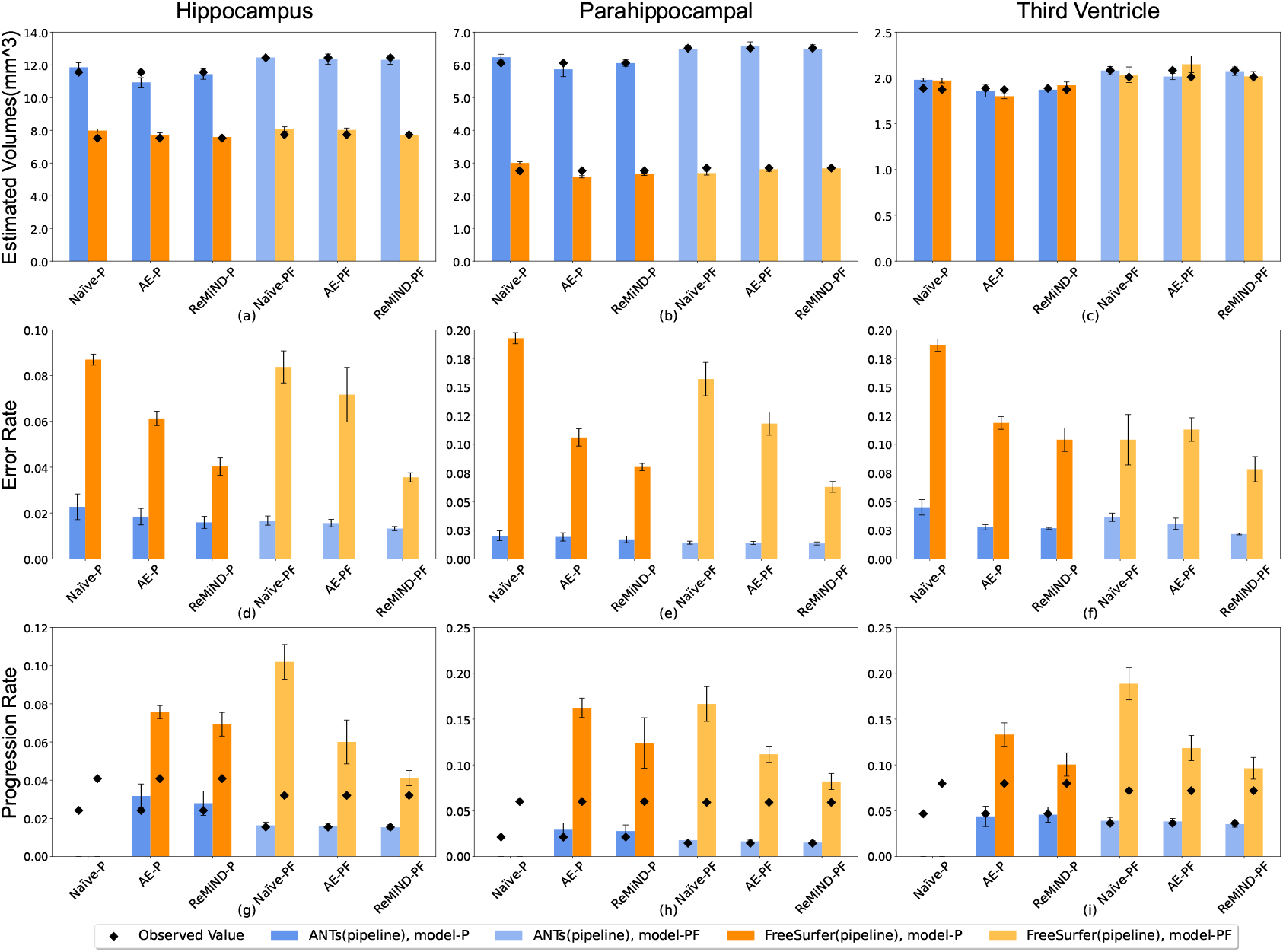
Comparison of volumetric features for three brain regions estimated on observed MRI images and MRI images generated from naive imputation, autoencoder, and ReMiND models. Results were averaged across 10 test sets. Error bars show standard errors across test sets. Estimated Volumes compare the observed volumes (represented as black diamond) and the volumes estimated from images generated with different models in panels a-c. Panels d-f present the comparison of error rates, where a lower value indicates better performance. In panels g-i, the progression rate is depicted using imputed images through bars, while the progression rate using observed images is represented by black diamonds. The volumes were estimated and compared with ANTs and FreeSurfer pipelines. P indicates the past image. PF indicates past and following images.

## 5. CONCLUSIONS

In this study, we introduced an innovative diffusion model-based framework for 3D longitudinal structural MRI imputation with an aim to generate missing 3D brain images at a specific single visit. To achieve this, the 3D image is partitioned into uniform local-continuous clips, with each clip consisting of three consecutive slices of the MRI image. Notably, our method distinguishes itself from conventional approaches by employing sets of 3D clips as input, thereby enhancing the proposed model’s ability to capture comprehensive global information from the entire MRI dataset during the training phase. The model utilizes single past or both past and following visits in the temporal direction to impute missing structural MRIs. Experimental results showed that our model consistently outperformed two comparator techniques: last image carried forward (i.e., forward filling) and imputation using an autoencoder. The comparison of similarity metrics highlighted our proposed model’s ability to accurately generate high-quality images for imputing missingness. We compared the volumes of a set of image derived phenotypes (IDPs) estimated from the generated and observed images using two different pipelines (ANTs and FreeSurfer). The relatively low error rates and accurately estimated rates of change in volume over time demonstrated that our proposed models can generate plausible whole-brain, 3D structural MRI data. Importantly, by imputing full 3D images rather than IDPs directly, researchers can flexibly utilize the imputed images in downstream statistical or predictive models, including using the generated images for further imputation of missing data in the temporal dimension. The proposed models alos has the potential to be beneficial for generating missing data in various other medical imaging contexts.

Across all experiments, the ReMiND-PF model, which conditioned on both past and future visits, outperformed the ReMiND-P model which solely relied on the past timepoint. This may be due to the ReMiND-PF model’s use of more information to generate images, highlighting the importance of quantity and quality of available information for imputing missingness. We evaluated the performance of the models on each clinical status group (CN, MCI, AD) and a combined all-status group and found that both ReMiND-P and ReMiND-PF performed well in all scenarios.

One limitation of our study lies in the utilization of either a single past image or both past and following images as conditional information for missingness imputation while ensuring a 6-month interval between consecutive visits. Future investigations could involve expanding the approach to incorporate multiple images with diverse visit interval timing. Additionally, there is potential for further research on downstream analyses, such as using the imputed imaging trajectories to develop models that predict the progression of Alzheimer’s disease.

## Data Availability

All data produced in the present work are contained in the manuscript

## Acknowledgments

Research reported in this publication was supported by the National Institutes of Health under award number R21AG075574. The content is solely the responsibility of the authors and does not necessarily represent the official views of the National Institutes of Health. Data collection and sharing for this project was funded by the Alzheimer’s Disease Neuroimaging Initiative (ADNI) (National Institutes of Health Grant U01 AG024904) and DOD ADNI (Department of Defense award number W81XWH-12-2-0012).

## Author Contributions

CY, RAH, and KAL conceived the essential concepts of the manuscript and directed the research. CY and JD contributed to data collection, data analysis, model development, and model evaluation. CY drafted the manuscript. NT, KX, RAH, and KAL supervised the implementation of the concept and provided critical revisions of the drafted manuscript. NT, RAH, and KAL obtained funding for the research. All authors gave final approval of the submitted manuscript.

## Conflict of Intetersts

Dr. Hubbard reports grant funding from Pfizer, Merck and Johnson & Johnson. The other authors report no conflicts of interest.

## Appendix A. Supplementary Material

**Table A.4.** The 28 regions on the cortical surface of the brain.

|  |  |
| --- | --- |
| Amygdala | Pallidum |
| Caudal Anterior Cingulate | Paracentral |
| Caudal Middle Frontal | Parahippocampal |
| Entorhinal | Pars Orbitalis |
| Fourth Ventricle | Pars Triangularis |
| Hippocampus | Posterior Cingulate |
| Inferior Lateral Ventricle | Rostral Anterior Cingulate |
| Inferior Parietal | Rostral Middle Frontal |
| Inferior temporal | Superior Frontal |
| Insula | Superior Parietal |
| Isthmus Cingulate | Superior Temporal |
| Lateral Ventricle | Thalamus |
| Medial Orbitofrontal | Third Ventricle |
| Middle Temporal | Transverse Temporal |

**Table A.5.**
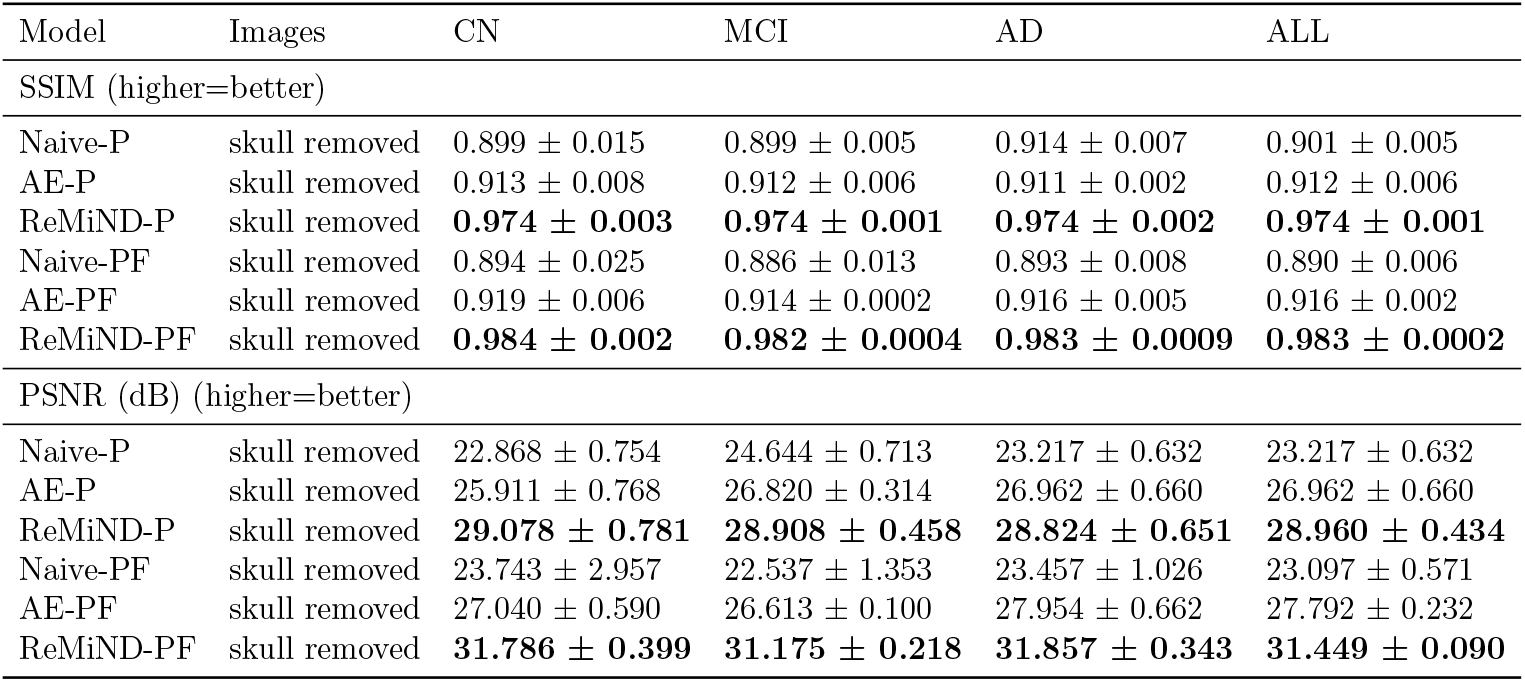
Comparison of model performance averaged across 10 test sets. Performance was measured with structural similarity index (SSIM) and peak signal-to-noise ratio (PSNR) in decibels (dB) on the generated MRI images with the skull removed (i.e., voxels within the group template brain mask only). Performance was evaluated overall and separately by clinical group (CN, MCI, and AD). P indicates the imputation method conditioned on the most recent past image. PF indicates the imputation method conditioned on both the most recent past image and the closest following image. AE indicates imputation using an autoencoder. Bold values indicate the best performing imputation method for a given clinical diagnosis group within the P or PF condition.

